# Symptomatic, Presymptomatic, and Asymptomatic Transmission of SARS-CoV-2

**DOI:** 10.1101/2021.07.08.21259871

**Authors:** Steven J. Krieg, Jennifer J. Schnur, Marie L. Miranda, Michael E. Pfrender, Nitesh V. Chawla

## Abstract

**Importance:** Asymptomatic and presymptomatic carriers of SARS-CoV-2 are an ongoing and significant risk for community spread of the virus, especially with the majority of the world still unvaccinated and new variants emerging.

**Objective:** To quantify the presence and effects of symptom presentation (or lack thereof) on the community transmission of SARS-CoV-2.

**Design:** A cohort of 12,960 young adults participated in health reporting, contact tracing, and COVID-19 testing for 103 days between August 10 and November 20, 2020.

**Setting:** A mid-sized university campus in Indiana, United States.

**Participants:** University students, most of whom are 18-23 years old (67%) and living in congregate on-campus housing (60%). Of the 12,960 students, 1,556 (12.0%) tested positive for COVID-19 during the 103 day period. Of the positive cases, 1,198 reported sufficient health check data (7 days prior and 7 days post diagnosis) to be classified as asymptomatic or symptomatic.

**Main Outcome:** Secondary attack rate, based on presentation or absence of symptoms and type of symptoms calculated with respect to confirmed close contacts and a 14-day incubation period, varies on the type of symptom, timing of symptoms, and absence of symptoms. A quantifiable understanding of SAR on the longitudinal data of more than one thousand subjects in a university environment provides keen insights about developing strategies to respond to the continued prevalence of COVID-19 in the unvaccinated world and growth of variants.

**Results:** 32.5% of all cases reported no symptoms within a 15-day window centered on their positive test (7 days prior, the day of the positive test, and 7 days after). The secondary attack rate (SAR) of asymptomatic COVID-19 index cases was 19.1%. The SAR of symptomatic index cases was 25.4%, and while the onset timing of symptoms did not affect transmission, the presence of certain symptoms like fever, shortness of breath, and dry cough increased the SAR as high as 30.0%.

**Conclusions and Relevance:** Asymptomatic rates of transmission of SARS-CoV-2 are much higher than has been estimated in prior studies and continue to pose a significant and ongoing risk in the pandemic, especially with the prevalence of variants like the Delta variant. In addition, different symptoms are associated with varying rates of transmission, posing a significant challenge in how to diagnose or assess risk through mechanisms such as daily health checks for symptom reporting, a practice commonly in place for entry into schools, offices, restaurants, etc. Given the uncertain nature of symptoms and varied transmission rates, this study suggests a broader embrace of masking, social distancing and testing might be needed to counter the variants until higher global vaccination rates can be achieved.

## Introduction

After the emergence of SARS-CoV-2 in Wuhan in late 2019, scientists quickly discovered the virus could be transmitted through asymptomatic carriers^1,2^. However, estimates for the percentage of asymptomatic cases vary from 40.6%-76.5%^3–7^. Differences across studies may result from variable cohort sizes; differences in demographics, social patterns, and community interaction; and an absence of data that is able to distinguish true asymptomatics—individuals who never experience symptoms—from presymptomatics—those who have no symptoms at the time of a positive test, but who later become symptomatic. Systematic review articles estimate that 30-45% of cases are asymptomatic, but also acknowledge that available case reports are limited and lacking in longitudinal data^8,9^. Some studies find that asymptomatic cases are less transmissible^10–13^, but researchers have yet to conclusively quantify the prevalence of asymptomatic cases and their role in the ongoing pandemic. For example, two meta-analyses have estimated secondary attack rates (SAR) of 18.0% and 21.1% for symptomatic cases and 0.7% and 1.9% for asymptomatic ones, but many of these reports have been limited by a lack of longitudinal data^10,14^. Our results suggest that these rates may be much higher than previously estimated for asymptomatic cases, as well as different symptoms if they are reported during a diagnosis.

To contribute to an understanding of the importance of asymptomatic and presymptomatic cases, as well as patterns of symptom presentation in confirmed infections, we present an analysis of symptom, testing, and contact tracing data collected during the Fall 2020 semester at a mid-sized Midwestern university. With a cohort of 12,960 students and 1,592 confirmed cases spread over 103 days, our analysis represents (to the best of our knowledge) the richest longitudinal study of its kind about asymptomatic and presymptomatic and symptomatic transmission of SARS-CoV-2. This study demonstrates that there is a varying SAR associated with different symptoms or even timing of emergence of different symptoms. This variability presents a significant challenge in successfully containing the pandemic, given the relatively low rates of global vaccination and threats of the variants such as the Delta variant^15^. In addition, as the economies opened up, places of work, education, and recreation embraced the idea of a daily health check as a mean to determine risk, which is also unreliable given the varying presentation and timing of symptoms (if any) and the associated SAR. This study suggests that until the population is sufficiently vaccinated, universal masking, broader testing, and social distancing might be the only reasonable mechanisms that can help overcome the risk of spread through asymptomatic, presymptomatic, or symptomatic transmission.

### COVID-19 and University Preparation

Along with the return to in-person classes in August 2020, the university initiated a series of measures to mitigate the risk of COVID-19 on campus. Of particular relevance to this analysis, the uUniversity implemented a daily self-reported health check, diagnostic testing, intensive contact tracing protocols, strict quarantine/isolation policies, and (roughly 11 days after the start of the fall semester) regular and required surveillance testing. Despite these efforts (detailed in the Methods section), the number of positive cases increased rapidly in the early days of the semester. Undergraduate students returned to campus during the first week of August, and classes began on August 10. After 151 students tested positive during the first week of the semester, the university halted in-person instruction and moved all classes to an online format. Graduate and professional students resumed in-person classes on August 24, while undergraduates remained online until September 2. This decision, along with other efforts, resulted in decreasing case numbers: while 601 students tested positive in August, the number of student cases decreased to 151 in September^16^. Positive test counts began to rise again in mid-October, and by the last day of the semester (November 20) a total of 1,561 students had tested positive on campus. Of the positive cases, we considered the self-reported symptom data for the 1,198 positive students who were at least 50% compliant with daily health checks within the 15-day window surrounding their positive test.

## Methods

### Symptom Reporting and COVID-19 Testing

Every day during the semester, each student on campus was asked to submit a brief electronic health survey inquiring if that student had experienced any of the 11 symptoms described in Table 1 or suspected they had been exposed to COVID-19. If a student reported exposure to a COVID-19 positive individual or a primary symptom (fever, shortness of breath, or loss of taste/smell), or reported any other symptoms for two consecutive days, they were administered a Sofia SARS Antigen Fluorescent Immunoassay (Quidel) rapid antigen test. If the antigen test yielded a negative result, the student was immediately administered a reverse transcription–polymerase chain reaction (PCR) test (performed by a local commercial laboratory primarily using a Roche platform) and temporarily quarantined until the PCR result was returned (usually 1-2 days). Of the individuals who tested negative via rapid antigen, we observed that 8.1% were false negatives and the same-day PCR test yielded a positive result.

**Table 1.** A summary of the symptoms monitored by daily health checks.

| Brief Description | Full description |
| --- | --- |
| Body aches/chills | Body aches and chills (not explained by exercise or cold environment) |
| Congestion | Persistent congestion or runny nose (not from a chronic or known condition) |
| Diarrhea | Diarrhea (more than 3 loose stools in 24 hours) |
| Fever | Fever of $\geq 100.4$ on two occasions 5 minutes apart |
| Headache | Headache (not from a chronic or known condition) |
| Nausea or vomiting | Nausea or vomiting |
| New dry cough | New dry cough (not from a chronic or known condition) |
| Loss of taste/smell | New loss of taste or smell |
| Shortness of breath | New shortness of breath or difficulty breathing (not from a chronic or known condition) |
| Sore throat | Painful sore throat (not from a chronic or known condition) |
| Congestion | Persistent congestion or runny nose (not from a chronic or known condition) |
| Fatigue | Unexplained Fatigue (not from lack of sleep) |

When a test yielded a positive result, the student was instructed to isolate for 14 days. A response team then attempted to identify anyone who had been in close contact with the student. Confirmed close contacts were instructed to quarantine and administered an antigen test. If the test produced a negative result, they were also administered a PCR test, instructed to quarantine, and called for additional tests on days four and seven after exposure. If all tests were negative by day seven, students were released from quarantine.

In addition to testing students who reported symptoms or exposure via close contact, the university also introduced surveillance testing to identify asymptomatic or presymptomatic cases. Every day, an additional sample of students was randomly selected and administered a PCR test in an on-campus facility (using the ThermoFisher TaqPath assay). In most cases the surveillance testing yielded negative results and no further action was taken; however, the positive testing students were instructed to isolate for 14 days. The sampling size and testing procedure varied over the course of the semester according to available testing resources and number of confirmed active cases. The number of surveillance tests administered increased over the course of the semester, so it is likely that some positive cases escaped detection, especially earlier in the semester.

### Data Preprocessing

From the symptom, testing, and contact tracing processes outlined above, we included data according to the following constraints:

- Subjects were active students (i.e. presently enrolled in an undergraduate, graduate, or professional degree program and registered for one or more courses) and residing in the local area. Although faculty and staff members were included in the testing and symptom reporting, their interactions and activities on campus may differ from students. Additionally, most of the positive cases were students.
- Subjects were at least 50% compliant with health checks within the 15-day window centered on their positive test (7 days prior, the day of the positive test, and 7 days after). Compliance is defined as the proportion of daily health checks completed within the relevant time frame surrounding a student’s positive test. We alternatively considered measuring student compliance across the whole semester, but found that this introduced several cases that were generally compliant but had many missing records near a positive test.
- The date of a positive test was between August 10 (the first day of the fall semester) and November 20 (the last day of the semester.

Health checks were more likely to be missing on Saturdays and Sundays than on other days. However, we are concerned with symptom onset relative to the date of a positive test, and positivity rates were relatively consistent throughout the week. We therefore chose to ignore missing health checks rather than risk introducing bias via imputation.

### Identifying Asymptomatics

We considered a subject to be an asymptomatic case if they met the aforementioned inclusion criteria and reported no symptoms within the 15-day window centered on their positive test (7 days prior, the day of the positive test, and 7 days after). If they reported any of the 11 symptoms once, we considered them to be symptomatic. We subcategorized symptomatic cases as presymptomatic (symptomatic only after positive test) if they did not report any symptoms in the seven days prior to a positive test or on the date a positive test was administered. For example, if a student reported a dry cough on August 10 and tested positive via rapid antigen on the same day, we consider them symptomatic but not presymptomatic.

### Analysis of Symptom Presentation

To compute the statistically significant windows for each symptom, we analyzed our sample of 1,198 cases against a sample of 1,198 negative cases. To generate the negative sample, for each date of the semester *d* we randomly selected *n* subjects such that *n* was equal to the number of positive tests produced on date *d*. Subjects were eligible as long as they:

1. Did not test positive at any point (even before August 10 or after November 20).
2. Were at least 50% compliant with health checks within 14 days of the given date (7 days prior, the current date *d*, and 7 days after). This is the same compliance criteria we used for including positive cases.
3. Had not already been included in a negative sample for another date in the semester.

This procedure produced a sample of non-positive students that contained sufficient symptom data and matched the daily distribution of positive cases. We then utilized Fisher’s exact test (one-sided) to test the null hypothesis that the proportion of positive cases reporting a given symptom on a given day was equal to the proportion of students from the negative sample who reported the symptom. We report as statistically significant any symptom and day combination with *p <* 0.05.

Figure 1 shows that three symptoms—congestion, headache, and dry cough—were statistically significant five days before a positive test. Fever and loss of taste/smell—two major symptoms of COVID-19—are not statistically significant until three and two days, respectively, before a positive test. Further, the representation of several symptoms does not peak until several days after a positive test; for example, loss of taste and smell is reported by the highest proportion of cases on day four.

**Figure 1.**
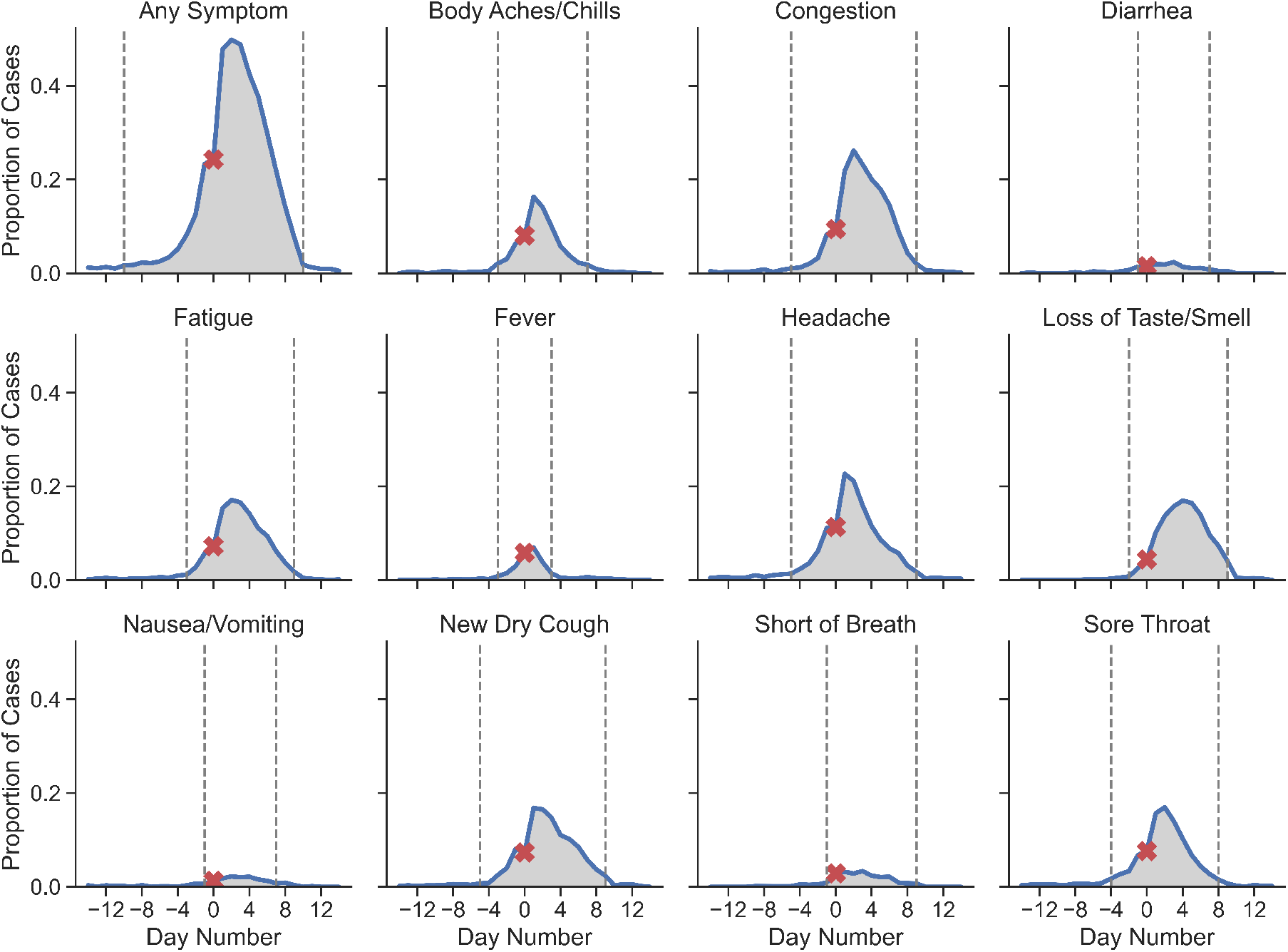
The reported symptoms from 14 days prior to 14 days after a positive test indicate that symptoms have different average onset times. Day 0 is the date of a positive test, marked by a red X. The shaded regions (bounded by vertical lines) indicate periods in which symptoms were statistically significant.

### Transmission and Secondary Attack Rate

The Secondary Attack Rate (SAR) is commonly used by epidemiologists to quantify the contagiousness of a disease, and is defined as the proportion of individuals who are infected after being exposed to a COVID-positive individual:

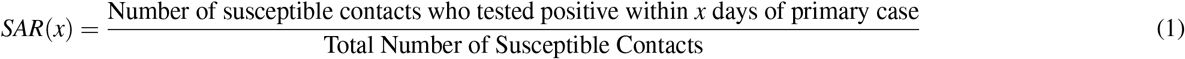

We define the incubation period *x* as the number of days between the index case and secondary case’s positive test. The dates associated with our contact tracing records were often noisy, missing, or unable to account for multiple occurrences of contact between pairs of students, so we used the primary case’s positive test date as an approximation for the exposure date. Household members generally serve as a proxy for susceptible contacts in computing SAR. However, we instead utilized our contact tracing observations, which is less likely to contain false positives (i.e. students who live at the same address but were not in close contact). Any false negatives (missing contacts) are excluded from both the numerator and denominator and are thus less impactful to the SAR calculation than false positives. We define a susceptible contact as any close contact who had not already tested positive for COVID-19 earlier in the semester. Close contacts were identified manually by contact tracers.

## Results

Of the 1,198 positive cases with symptom data, 389 (32.5%) reported no symptoms within the 15-day window centered on their positive test and thus appear asymptomatic (Table 2). An additional 485 cases (40.5%) reported symptoms prior to or on the same day as their positive test, whom we classify as symptomatic. An additional 324 (27.0%) cases reported symptoms subsequent to a positive test and were thus presymptomatic at the time of the test.

**Table 2.** Summary of COVID-19 data collected during the Fall 2020 semester (Aug. 10–Nov. 20).

| Description | Count |
| --- | --- |
| Active students | 12,960 |
| Daily health checks submitted | 868,196 |
| COVID-19 tests | 73,950 |
| Close contacts identified | 3,736 |
| Total positive cases | 1,556 |
| Positive cases included in study | 1,198 |
| Symptomatic | 809 (67.5%) |
| Symptomatic (before positive test) | 485 (40.5%) |
| Symptomatic (only after positive test) | 324 (27.0%) |
| Asymptomatic | 389 (32.5%) |

Taking into account a 14-day potential transmission window, Figure 2 shows the computed SARs for asymptomatic, symptomatic, and presymptomatic cases. Note that these measurements are grouped by the index case’s symptomatic status; for example, any students who were exposed via contact with an asymptomatic case only contribute to the SAR for asymptomatic cases, regardless of whether they (the secondary cases) become symptomatic. At day 14 in the incubation period, the SAR was 19.1% for asymptomatic index cases, as compared to 25.% for all symptomatic index cases. In the first four days of the incubation period, the SAR was highest for symptomatic index cases, followed by presymptomatic index cases – and lowest for asymptomatic index cases. After day four, the SAR for presymptomatic exceeds the SAR for symptomatic index cases, but the two values become mostly coincident. The higher initial SAR for symptomatic index cases is likely because they are testing positive later in their infections and therefore had more time to infect others before being isolated. Importantly, all three SARs become relatively flat after about day seven of the incubation period, which has implications for how long contact traced individuals need to remain in quarantine. These results are consistent with the choices of some local health departments to release individuals from quarantine after day 10 or after day seven with a negative test result.

**Figure 2.**
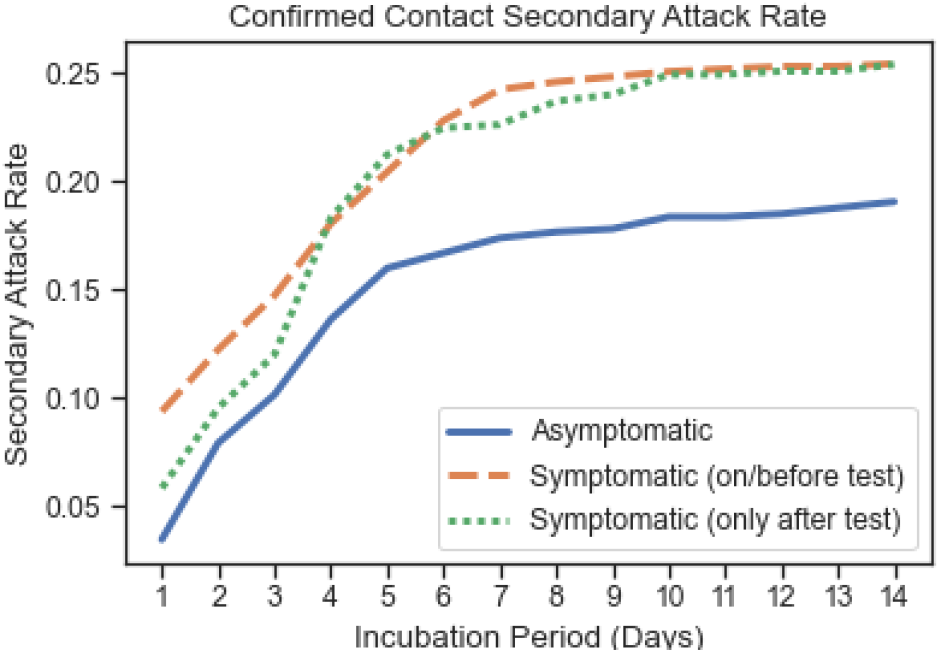
Observed secondary attack rates calculated with respect to symptomatic presentation of the index cases.

### Symptomatic Presentation and Transmission

Of the 1,198 positive cases with symptom data, we identified 807 (67.4%) symptomatic cases while observing significant variety in the timing and symptoms reported by these individuals. For example, the most-reported symptoms—headache and congestion—were reported by 38.8% and 35.6% of the 1,198 cases, respectively. All other symptoms were reported in fewer than 28% of cases. Additionally, the most correlated pair of symptoms—fatigue and body aches/chills—only produced a correlation coefficient of 0.41. Additionally, when we consider each symptom individually, we observe different average onset times and durations (see Supplementary Information). Importantly, 75.2% of positive cases reported experiencing no symptoms on the day of their positive test. This number decreases to 50.2% two days after a positive test. Thus, among the total cases, many individuals were presymptomatic at time of testing, which is relevant for designing surveillance testing programs.

We further observed that the presence or absence of individual symptoms did affect transmission. Figure 3 shows the secondary attack rate between confirmed close contacts based on the presence or absence of individual symptoms reported by the index cases. For some symptoms like fever, new dry cough, and shortness of breath, the difference in SAR was substantial and presented immediately. For body aches/chills, the difference in SAR was also substantial but did not present until about day four of the incubation period. For all four of these symptoms associated with higher SARs, the attack rate stabilized at roughly 30.0%, compared to the average SAR of 25.4% from Figure 2. Additionally, we found that the SAR for index cases that reported loss of taste/smell or diarrhea was in fact lower than for those that did not. While it would be tempting to suggest that individuals who experienced loss of taste/smell were more likely to maintain physical distancing or practice caution (perhaps because anosmia is such a distinct symptom of COVID-19), this would not fit our observed data. An alternative hypothesis is that the presence of certain symptoms, such as fever and shortness of breath, are correlated with higher viral load, which in turn leads to greater transmissibility. Future research on this topic could yield a more conclusive result.

**Figure 3.**
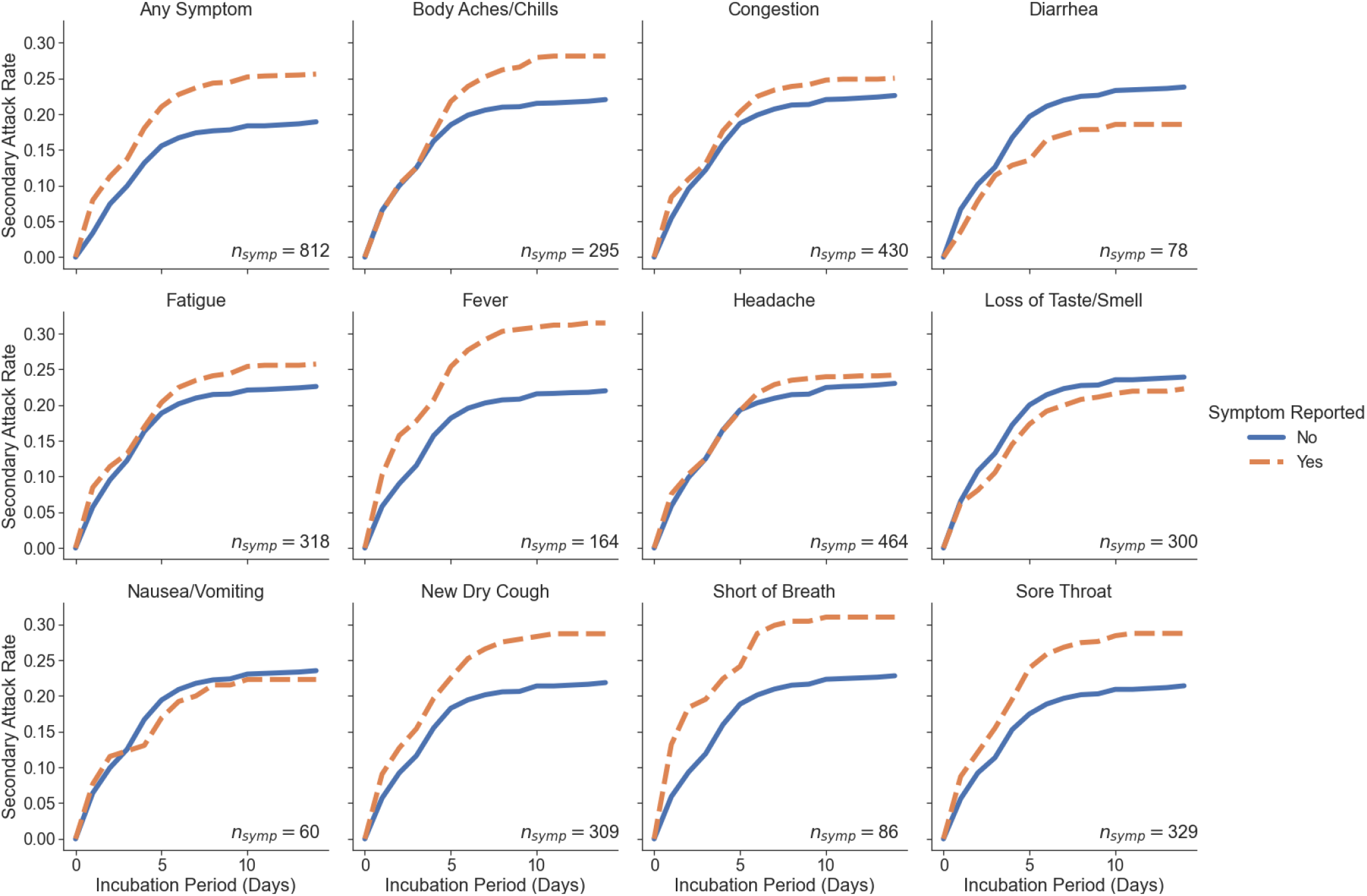
Secondary attack rates between confirmed contacts based on the presence or absence of each reported symptom. *n*_*symp*_ is the number of positive cases that reported the symptom at least once in the 14-day window surrounding their positive test.

## Discussion

Our key findings that 32.5% of confirmed cases were completely asymptomatic, and that asymptomatic cases were only 25% less reproductive than symptomatic cases suggests that prior studies have significantly underestimated the transmissibility of asymptomatic COVID-19 cases^10^. Until we achieve a broader vaccination coverage, asymptomatic transmission may remain a significant and invisible threat. With respect to symptom presentation, some symptoms such as congestion and headache were early onset and other symptoms like shortness of breath and loss of taste and smell were later onset, demonstrating that individuals who are experiencing even mild symptoms should exercise extreme caution. Moreover, this also suggests different symptoms are of different import, and daily health checks might not tell sufficient details about probability of transmission. We additionally observed that the presence of certain symptoms such as a fever or a dry cough increased the likelihood of transmission between close contacts by as much as 10%. These results suggest, resources permitting, that anyone exhibiting fever, body aches/chills, new dry cough, shortness of breath, or a sore throat should be tested immediately for COVID-19.

The secondary attack rate (SAR) was generally higher if the individual was symptomatic, except in cases where the symptom was diarrhea, nausea/vomiting, or loss of taste/smell, a commonly cited indicative symptom of COVID-19. Even the most frequently reported symptoms (e.g. headache, congestion) among confirmed positive individuals occurred in less than half of COVID-19 hosts. The breadth and variety of COVID 19 symptom presentation, paired with the variable timing of symptom onset, will continue to complicate our ability to identify and predict definitively who is infected with the virus. This analysis suggests that daily health checks that rely on symptom reporting for places of work, study, recreation, etc., might be misleading, and thus adherence to universal masking as a deterrent to transmission might be the stop-gap measure till better vaccination rates are achieved, globally. These varying SAR across presence or absence of symptom(s) becomes even more profound a challenge with the emergence of highly transmissible variants, such as the Delta variant.

While this study shows significant findings associated with longitudinal symptom presentation and SARS-CoV-2 contagion, it is not without limits. First, the cohort of positive cases is relatively homogeneous; it is composed entirely of university students, most of whom are undergraduates (92%), white (74%), 18-23 years old (67%), and many of whom (60%) are living in congregate on-campus housing. Clearly, our sample is not representative of the general population. Second, our analysis assumes a closed community. While many positive cases were traced to connections within the university, we cannot account for any transmission that may have occurred within the local community. Thus our estimates may represent a lower bound on secondary attack rates, but are likely good estimates for proportion of asymptomatic cases among young adults. A third limitation relates to the fact that contact tracing records are dependent on self-reporting of confirmed contacts from students who test positive. While missing data does not directly impact our SAR calculations, it runs the risk of introducing statistical bias. Fourth, our symptom analysis relies on accurate and consistent self-reporting. Although we discarded data from students who were less than 50% compliant with health check completion around the time of their positive test, such a threshold is arbitrary and cannot fully address other problems such as false negatives.

This study quantitatively establishes the role of asymptomatic and presymptomatic SARS-CoV-2 transmission in a university context. Our results suggest that asymptomatic and presymptomatic transmission has played a significant role in the ongoing pandemic and continues to pose a significant threat, especially in the context of emerging variants. As schools, restaurants, and other social institutions reopen, continued masking, social distancing, and widespread testing will remain essential until we can achieve high global vaccination rates.

## Data Availability

The data and code for this study are available from the corresponding author upon reasonable request.

## Acknowledgements

This study was sponsored by the Lucy Family Institute for Data and Society, which conceived of the study and the decision to submit the manuscript for publication.

## Author Contributions

S.J.K, J.J.S, M.L.M, and N.V.C conceived of the study. S.J.K. and J.J.S. modeled the data. S.J.K, J.J.S, and N.V.C performed the analysis. All authors reviewed and wrote the manuscript.

